# Longitudinal changes in dose-normalized psychotropic medication exposure around transfer to a specialized older adult mental health service: a retrospective longitudinal study in Peru

**DOI:** 10.64898/2026.09.15.26363167

**Authors:** Luis Macedo-Orrego, Paulo Ruiz-Grosso, Sonia Zevallos-Bustamante

## Abstract

**Objectives:** This research aimed to characterize longitudinal changes in dose-normalized psychotropic exposure around transfer to a specialized older adult mental health service in Peru, with the primary contrast comparing the last eligible pre-transfer consultation (T0) with six months after transfer. We additionally explored characteristics associated with average medication-score levels and potential modification of the T0-to-6-month change.

**Methods:** A retrospective longitudinal single-cohort study was conducted using routine clinical records from the Instituto Nacional de Salud Mental Honorio Delgado-Hideyo Noguchi in Lima, Peru. The primary analytic cohort comprised patients transferred to the older adult service who had valid longitudinal medication data at or before transfer and after transfer. A dose-normalized psychotropic medication score was calculated at approximately -6 months, -3 months, T0 (the last eligible consultation before transfer), +3 months, and +6 months. Longitudinal change was estimated using linear mixed-effects models with categorical time and participant-specific random intercepts; the prespecified primary contrast compared +6 months with T0. Sensitivity analyses evaluated alternative score definitions, age restrictions, influential observations, and participant-level cluster bootstrap resampling.

**Results:** The primary analytic cohort included 213 participants; mean age at T0 was 65.5 years (SD 7.2), 59.2% were female, and the most frequent diagnostic categories were psychotic disorders (52.1%) and depressive disorders (29.1%). The medication score was 17.87 points higher at +6 months than at T0 (95% CI 12.18 to 23.57; p < 0.001), and the exploratory adjusted estimate was similar (17.82 points; 95% CI 12.10 to 23.54; p < 0.001). The primary contrast remained positive across alternative score definitions, age-restricted analyses, influence analyses, and participant-level bootstrap resampling. In exploratory effect-modification analyses, each 10-year increase in age was associated with a 10.94-point greater T0-to-+6-month change (95% CI 2.33 to 19.55; p = 0.013), although this association did not remain statistically significant after false-discovery-rate correction (FDR-adjusted p = 0.091); no other tested modifier showed evidence of interaction after FDR correction.

**Conclusions:** Dose-normalized prescribed psychotropic exposure increased from a relative pre-transfer low point at T0 to 6 months after transfer, with similar estimates across sensitivity analyses. In the absence of a contemporaneous comparison group, this finding should be interpreted as a temporal association around service transfer rather than evidence that transfer itself caused the increase or that the resulting prescribing was clinically beneficial, harmful, or inappropriate.

## Introduction

Population aging is accelerating worldwide [1]. In Peru, the health and quality-of-life implications of population aging have been recognized for years [2], and demographic analyses project continued growth of older age groups [3]. In the Americas, life expectancy at age 65 increased from 17.1 years in 1990 to 19.2 years in 2019, whereas healthy life expectancy increased from 12.2 to 13.6 years over the same period [4]. National reports estimate that adults aged 60 years or older now represent approximately 14.3% of the population in Peru [5]. As more people live to older ages with chronic physical and mental disorders, the need for long-term pharmacological treatment is likely to grow, making periodic medication review increasingly relevant to both clinical care and health systems.

Older adults receiving mental health care often have multimorbidity, frailty, cognitive impairment, and other age-related vulnerabilities that may complicate long-term pharmacological treatment [6]. This population also presents pharmacokinetic and pharmacodynamic changes, including alterations in hepatic and renal function, that may modify psychotropic exposure, response, and tolerability [7]. At the same time, mental disorders, whether long-standing or of more recent onset, affect a substantial proportion of older adults and may require psychotropic treatment. For example, a meta-analysis of 31 studies from 17 Latin American and Caribbean countries estimated a pooled all-cause dementia prevalence of 10.66% [8], while the 10/66 study of 5,886 adults aged 65 years or older in Peru, Mexico, and Venezuela reported depressive episode prevalence ranging from 4.5% to 5.1% across sites [9]. However, evidence regarding the efficacy, safety, and optimal dosing of several psychotropic medications remains less extensive in older than in younger adult populations [10]. Taken together, these considerations support periodic reassessment of treatment indication, dose, medication combinations, and treatment duration in geriatric mental health care [6,6,10].

Epidemiological evidence shows that psychotropic medication exposure is common in later life, although estimates vary by population, setting, medication class, and exposure definition. In a recent nationwide Australian study, 31.1% of adults aged 65 years or older received at least one psychotropic medication during a three-month period [11], and studies from New Zealand and Canada have likewise documented substantial and changing psychotropic medication use among older adults over time [12,12]. Potentially inappropriate prescribing remains an important concern in older populations [14], and psychotropic medication use has been associated with fall-related injuries, hospitalization, and mortality; for example, in a nationwide Swedish study of 1,288,875 adults aged 65 years or older, antidepressant use was associated with higher adjusted odds of fall injury (OR 1.42, 95% CI 1.38 to 1.45) [15]. These findings reinforce the need to characterize psychotropic exposure carefully while distinguishing medication burden from prescribing appropriateness.

Medication count and dose intensity capture different aspects of psychotropic exposure. In established psychiatric populations, older adults may receive a similar number of psychotropic medications as younger adults while being prescribed lower doses of specific agents [16]. In a European dataset of 32,062 inpatients with schizophrenia, antipsychotic doses decreased with advancing age, more steeply after approximately 55 years; only one of eight schizophrenia guidelines reviewed systematically addressed specific aspects of pharmacotherapy in older adults [17]. Dose-standardization approaches such as defined daily doses, chlorpromazine equivalents, and percentages of a reference maximum dose have been used to compare treatment intensity across antipsychotic agents [18]. However, systematic comparison of these methods shows that they can yield different equivalence estimates and that no single method constitutes a universal gold standard [19]; moreover, defined daily doses were developed primarily as standardized measures of drug consumption and may be misleading when interpreted as pharmacological dose equivalence [20]. These differences underscore the importance of explicitly defining the dose-standardization method used to compare prescribing intensity across agents.

Latin American evidence illustrates the heterogeneity of psychotropic exposure across settings. In a cohort of 63,857 adults aged 60 years or older in Argentina, central nervous system-active polypharmacy was present in 7.1%, with particularly strong associations for schizophrenia (OR 7.93, 95% CI 4.64 to 13.56) and bipolar disorder (OR 7.20, 95% CI 5.45 to 9.50) [21]. In Peru, a retrospective study of 430 outpatients with schizophrenia at a specialized mental health institution found that 84.4% received more than one psychotropic medication and 40.5% received more than one antipsychotic [22]. In a community-pharmacy study of 158 older adults in Trujillo, 93.7% had at least one STOPP indicator and 53.8% at least one START indicator [23]. These regional studies have primarily characterized medication prevalence, polypharmacy, or prescribing appropriateness rather than longitudinal change in dose-normalized psychotropic exposure.

Longitudinal studies indicate that psychotropic prescribing can change substantially around late-life care transitions, although the direction of change is not uniform. In a Norwegian nursing-home cohort, the incidence of each major psychotropic drug category was highest during the first six months after admission, reaching 49.4% for antipsychotics [24]. In Northern Ireland, antipsychotic dispensing increased from 8.2% before entry to 18.6% after entry into care, while hypnotic dispensing increased from 14.8% to 26.3% [25]; a national Australian cohort of 322,120 residents likewise documented marked changes in psychotropic dispensing around residential-care entry [26]. Conversely, a systematic review of 19 specialist outpatient studies including 10,914 participants found four randomized trials reporting statistically significant reductions in medication load, although all included studies were judged at high risk of bias [27]. Existing work has therefore focused largely on residential or nursing-home transitions, dispensing prevalence, medication counts, individual drug classes, or prescribing appropriateness. Evidence describing longitudinal change in an operational, dose-normalized measure of psychotropic exposure around transfer to a specialized older adult outpatient mental health service appears limited, particularly in Latin America. We therefore aimed to characterize longitudinal changes in dose-normalized psychotropic exposure around transfer to the older adult service of the Instituto Nacional de Salud Mental Honorio Delgado-Hideyo Noguchi between 2022 and 2024, with the prespecified primary contrast comparing the last eligible pre-transfer consultation (T0) with six months after transfer. We additionally explored characteristics associated with average medication-score levels and whether selected patient characteristics modified the T0-to-6-month change.

## Materials and Methods

### Study design and setting

A retrospective longitudinal single-cohort study was conducted using routinely collected clinical records from the older adult service of the DEIDAE de Adultos y Adultos Mayores at the Instituto Nacional de Salud Mental Honorio Delgado-Hideyo Noguchi (INSM), Lima, Peru. The study included patients whose follow-up in the older adult service began in the period between 1 January 2022 and 31 December 2024.

The INSM is a specialized referral center for mental health care that receives patients that might be referred from primary care facilities, community mental health centers (CMHCs) and general hospitals. It operates within the Seguro integral de Salud (SIS), which is a largely tax-financed scheme serving poor and vulnerable populations, entrepreneurs, and self-employed workers; while EsSalud is a contributory scheme that primarily covers salaried formal-sector workers and their families; in 2023, these systems covered approximately 62% and 26% of the Peruvian population, respectively [28]. This distinction is particularly relevant in Peru, where 70.9% of the employed population worked in informal employment in 2024 [29]. Among adults aged 60 years or older, 54.3% were affiliated with SIS and 34.8% with EsSalud in the fourth quarter of 2024 [30]. Also, Peru’s mental health reform has progressively shifted service delivery from a predominantly hospital-centered model toward community-based care, while specialized mental health institutes continue to play referral, training, and clinical-support roles [31]. Consequently, some patients included in this study had initiated treatment at the INSM many years before reaching older adulthood and were subsequently transferred to the older adult service. Thus, for a proportion of the cohort, transfer to the older adult service represented a transition between services within the same institution rather than the initiation of specialized mental health care.

Time was structured around transfer to the older adult service. T0 was defined as the last eligible clinical consultation before transfer. Medication information was obtained from the eligible consultation closest to each prespecified time point: approximately 6 months before transfer (-6), 3 months before transfer (-3), T0, 3 months after transfer (+3), and 6 months after transfer (+6). Because visits occurred as part of routine clinical care, measurement times were approximate and participants might not contribute observations at every time point.

### Participants

The source population consisted of patients who were transferred to and received outpatient psychiatric care in the older adult service of the INSM during the study period. Potentially eligible patients were identified from institutional clinical records. Because all patients meeting the eligibility criteria were intended to be included, no probabilistic sampling was performed. For the primary longitudinal analysis, participants were required to have documented transfer to the service, at least one valid medication-score observation at or before T0, and at least one valid observation after T0.

The original protocol included an age-based operational eligibility criterion at the nominal start of follow-up, 6 months before transfer. In routine clinical practice, however, assignment to the older adult service is not determined exclusively by chronological age, and some patients may be transferred before age 60 according to clinical and service-level considerations. The primary analytic cohort was therefore defined by actual transfer to the service and longitudinal medication-data availability rather than by a strict age threshold. Robustness to the protocol wording was evaluated in sensitivity analyses restricting the cohort to participants aged 60 years or older at T0, 60 years or older at the nominal start of follow-up, and older than 60 years at the nominal start of follow-up. Consultations consisting only of prescription renewal, administrative records, or missed appointments were not considered eligible clinical visits for medication abstraction.

### Outcome: psychotropic medication score

The primary outcome was a continuous psychotropic medication score calculated at each eligible visit. For each scheduled psychotropic medication, the prescribed total daily dose was divided by a prespecified medication- and formulation-specific reference upper daily dose and multiplied by 100; contributions were then summed across medications prescribed at that visit. The protocol term ‘maximum recommended daily dose’ was operationalized during analytic reconstruction as this reference upper dose because a single universal maximum is not defined consistently across medications, formulations, and sources. Prescriptions documented as PRN-only were not added to the scheduled daily-dose score. Visits explicitly documenting no psychotropic medication were assigned a score of zero, whereas blank or explicitly unknown medication information was treated as missing rather than as zero. An alternative reference-dose specification was evaluated in sensitivity analysis. Medication- and formulation-specific primary reference upper doses, together with the alternative values used in sensitivity analysis, are provided in Supplementary Table S2.

The score was treated as an operational dose-based indicator of psychotropic medication exposure. It does not establish dose equivalence across drug classes and should not be interpreted as a validated measure of prescribing appropriateness, anticholinergic burden, adverse-effect burden, toxicity, or overmedication. Accordingly, a higher score indicates greater dose-normalized psychotropic exposure but not necessarily inappropriate treatment.

### Exposure and time variable

The main independent variable was time relative to transfer to the older adult service, modeled as a categorical variable with five levels: -6 months, -3 months, T0, +3 months, and +6 months. T0 was the reference category in the primary mixed-effects model. The prespecified primary longitudinal estimand was the model-estimated mean difference in medication score between +6 months and T0. The remaining time contrasts were secondary descriptions of the trajectory around transfer.

### Covariates

Sociodemographic variables included age at T0, sex, attained education, and marital status. Clinical variables included psychiatric diagnostic categories coded from routine clinical records using ICD-10, diagnostic burden, duration of psychiatric illness, and duration of care at the INSM. Diagnostic categories were described broadly in Table 1. Candidate covariates evaluated in exploratory mixed-effects models were sex, age (scaled per 10-year increase), education, marital status, depressive disorder, bipolar disorder, psychotic disorder, anxiety or stress-related disorder, dementia, diagnostic burden (0-1, 2, or >=3 diagnostic categories), duration of psychiatric illness, and duration of care at the INSM.

**Table 1.** Baseline demographic and clinical characteristics.

| Variables | n (%) |
| --- | --- |
| **Sociodemographic characteristics** |  |
| Female sex | 126 (59.2%) |
| Age at T0, years, mean (SD) | 65.5 (7.2) |
| **Education** |  |
| Less than complete primary | 26 (12.3%) |
| Complete primary | 79 (37.3%) |
| Complete secondary | 77 (36.3%) |
| Higher education | 30 (14.2%) |
| **Marital status** |  |
| Single | 109 (51.2%) |
| Married | 64 (30.0%) |
| Cohabiting | 17 (8.0%) |
| Widowed / separated / divorced | 23 (10.8%) |
| **Clinical history** |  |
| Duration of psychiatric illness, years, mean (SD) | 23.3 (16.2) |
| Duration of care at INSM, years, mean (SD) | 13.8 (13.1) |
| **Psychiatric diagnoses at T0** |  |
| Psychotic disorders | 111 (52.1%) |
| Depressive disorders | 62 (29.1%) |
| Anxiety/stress-related disorders | 27 (12.7%) |
| Dementia | 23 (10.8%) |
| Organic mental disorders | 18 (8.5%) |
| Bipolar disorders | 18 (8.5%) |
| Substance-related disorders | 9 (4.2%) |
| Intellectual disability | 4 (1.9%) |
| Personality disorders | 3 (1.4%) |
| Somatoform disorders | 3 (1.4%) |
| Insomnia | 1 (0.5%) |
| **Diagnostic burden at T0** |  |
| 0-1 diagnostic categories | 154 (72.3%) |
| 2 diagnostic categories | 46 (21.6%) |
| >=3 diagnostic categories | 13 (6.1%) |

Notes: values are n (%) unless otherwise specified. Education percentages use non-missing observations (n = 212). Duration of psychiatric illness is based on 195 participants; other listed baseline characteristics use n = 213 unless otherwise specified. Psychiatric diagnostic categories are not mutually exclusive. T0 denotes the last eligible clinical consultation before transfer.

### Statistical analysis

Continuous variables and observed medication scores were summarized using means and standard deviations, while categorical variables were described using frequencies and percentages. The primary longitudinal analysis used a linear mixed-effects model with medication score as the outcome, categorical time as a fixed effect, and a participant-specific random intercept. The model was fitted by restricted maximum likelihood and used all available eligible repeated observations. T0 was the temporal reference. The overall effect of time was assessed with a Type III F test using Satterthwaite-approximated degrees of freedom. The prespecified +6-month versus T0 contrast was reported with a 95% confidence interval and two-sided p value without multiplicity adjustment because it represented the primary contrast.

Exploratory covariate screening was conducted using individual mixed-effects models that included time and one candidate covariate at a time, with the same participant-specific random intercept. For categorical covariates with more than two levels, the overall association of the covariate with medication score was assessed using a Type III Wald F test with Satterthwaite-approximated degrees of freedom. Candidate covariates with an overall p value <0.20 in the current analytic dataset were entered into the multivariable model. No time-by-covariate interactions were included in this model; therefore, covariate coefficients represent average differences in medication-score level across observed time points and not modification of the T0-to-+6-month change. The multivariable model used complete cases for the selected covariates, and no missing-data imputation was performed.

Separately, exploratory effect-modification analyses evaluated whether the prespecified T0-to-+6-month change differed according to sex, age, education, depressive disorder, anxiety or stress-related disorder, psychosis, or diagnostic burden. Psychosis was included a priori on clinical and theoretical grounds, independently of the p < 0.20 screening rule used for the Table 3 multivariable model. These analyses were restricted to T0 and +6 months and used a common complete-case dataset. Each model contained time, the candidate effect modifier, the main effects of sex, age, education, depressive disorder, anxiety or stress-related disorder, psychosis, and diagnostic burden, a single time-by-modifier interaction, and a participant-specific random intercept. Global interaction tests used Type III Wald F tests with Satterthwaite-approximated degrees of freedom. To address multiplicity across the seven global interaction hypotheses, Benjamini-Hochberg false-discovery-rate correction was applied. Age was modeled per 10-year increase. These interaction analyses were considered exploratory.

Estimated marginal means were obtained for each time point. Secondary pairwise comparisons across all time points were calculated with Tukey adjustment for multiple comparisons and are reported in Supplementary Table S3; these comparisons were considered supplementary to the prespecified primary contrast. Sensitivity analyses recalculated the primary contrast using the alternative medication-score definition, alternative age restrictions, and a diagnostic influence analysis excluding the 10 participants with the most extreme residual patterns. Model assumptions were assessed visually using residual-versus-fitted plots and quantile-quantile plots of conditional residuals and participant random intercepts.

To evaluate robustness to distributional departures and within-participant dependence, a nonparametric participant-level cluster bootstrap resampled participants with replacement and refitted the full five-time-point mixed-effects model in 2,000 bootstrap samples. The percentile bootstrap 95% confidence interval was reported. Because the covariate-adjusted analyses, effect-modification analyses, and sensitivity analyses were exploratory, interpretation emphasized effect size, confidence intervals, direction, and consistency rather than statistical significance alone. Covariate-specific coefficients were interpreted as associations rather than independent causal effects.

Analyses were performed in R version 4.5.2 (R Foundation for Statistical Computing, Vienna, Austria). Linear mixed-effects models were fitted with lme4 version 2.0.6 and lmerTest version 3.2.1; estimated marginal means and contrasts were obtained with emmeans version 2.0.4, with pbkrtest version 0.5.5 used where Kenward-Roger degrees-of-freedom calculations were required.

### Ethics

This study was based on retrospective information abstracted from routine clinical records and did not involve any direct contact with patients. The protocol (registration code 589-2024) was submitted to the Institutional Research Ethics Committee (Comité Institucional de Ética en Investigación, CIEI) of the Instituto Nacional de Salud Mental Honorio Delgado–Hideyo Noguchi (INSM) on July 8, 2024. The CIEI issued Exemption Certificate No. 001-2024-CIEI-INSM-"HD-HN" on August 8, 2024, documenting that the protocol was exempt from review by the CIEI. The exemption certificate did not include a separate statement regarding waiver of individual informed consent. The analytic dataset was de-identified, and a separate identification file was maintained only for data quality control and record linkage during data abstraction, with each participant assigned an alphanumeric code; access to identifiable information was restricted to authorized study personnel throughout the process.

## Results

### Participant characteristics

We included 213 participants in the primary analytic cohort. Mean age at T0 was 65.5 years (SD 7.2), and 126 participants (59.2%) were female. Among the 212 participants with available education data, 26 (12.3%) had less than complete primary education, 79 (37.3%) had complete primary education, 77 (36.3%) had complete secondary education, and 30 (14.2%) had higher education. Regarding marital status, 109 participants (51.2%) were single, 64 (30.0%) were married, 17 (8.0%) were cohabiting, and 23 (10.8%) were widowed, separated, or divorced (Table 1).

Psychotic disorders were the most frequent psychiatric diagnostic category (n = 111; 52.1%), followed by depressive disorders (n = 62; 29.1%), anxiety or stress-related disorders (n = 27; 12.7%), dementia (n = 23; 10.8%), organic mental disorders (n = 18; 8.5%), and bipolar disorders (n = 18; 8.5%). Mean duration of psychiatric illness was 23.3 years (SD 16.2) among 195 participants with available data, and mean duration of care at the INSM was 13.8 years (SD 13.1). At T0, 154 participants (72.3%) had 0-1 psychiatric diagnostic categories, 46 (21.6%) had two, and 13 (6.1%) had three or more (Table 1).

### Changes in medication score over time

Observed mean medication scores were 108.5 (SD 72.4) at -6 months, 105.6 (SD 71.8) at -3 months, 94.7 (SD 64.0) at T0, 106.0 (SD 66.1) at +3 months, and 114.4 (SD 68.6) at +6 months. The corresponding estimated marginal means from the primary linear mixed-effects model were 104.9, 101.6, 94.8, 106.1, and 112.7, respectively. There was an overall effect of time (F[4, 670.37] = 10.07; p < 0.001). For the prespecified primary contrast, the medication score at +6 months was 17.87 points higher than at T0 (95% CI 12.18 to 23.57; p < 0.001) (Table 2).

**Table 2.** Longitudinal medication-score trajectory.

| Timepoint | N | Observed mean (SD) | Estimated marginal mean | 95% CI |
| --- | --- | --- | --- | --- |
| -6 months | 124 | 108.5 (72.4) | 104.9 | 95.0-114.7 |
| -3 months | 143 | 105.6 (71.8) | 101.6 | 92.0-111.2 |
| T0 | 211 | 94.7 (64.0) | 94.8 | 85.7-103.9 |
| +3 months | 207 | 106.0 (66.1) | 106.1 | 97.0-115.2 |
| +6 months | 198 | 114.4 (68.6) | 112.7 | 103.5-121.9 |
| **Primary mixed-effects model** |  |  |  |  |
| **Test** | **Result** | **P value** |  |  |
| Global effect of time | F(4, 670.37) = 10.07 | <0.001 |  |  |
| Primary contrast: +6 months minus T0 | 17.87 (95% CI 12.18-23.57) | <0.001 |  |  |

Notes: observed values are arithmetic mean (SD). Estimated marginal means and the global time test are from the primary linear mixed-effects model with time as a categorical fixed effect and a participant-specific random intercept. T0 denotes the last eligible clinical consultation before transfer. The prespecified primary contrast is +6 months minus T0.

With T0 as the reference time point, medication scores were also higher at +3 months (beta = 11.27; 95% CI 5.66 to 16.88; p < 0.001), while scores at -6 months (beta = 10.06; 95% CI 3.35 to 16.78; p = 0.003) and -3 months (beta = 6.80; 95% CI 0.42 to 13.18; p = 0.037) were likewise higher than at T0. In the exploratory multivariable model, the +6-month versus T0 estimate was essentially unchanged (beta = 17.82; 95% CI 12.10 to 23.54; p < 0.001) (Table 3). Tukey-adjusted pairwise comparisons across all time points are provided in Supplementary Table S3.

**Table 3.** Individual and adjusted mixed-effects models for medication score.

| Variable | Individual<br>beta | 95% CI | P<br>value | Adjusted<br>beta | 95% CI | P<br>value |
| --- | --- | --- | --- | --- | --- | --- |
| **Time** |  |  |  |  |  |  |
| -6 vs T0 | 10.06 | 3.35-16.78 | 0.003 | 10.03 | 3.27-16.79 | 0.004 |
| -3 vs T0 | 6.80 | 0.42-13.18 | 0.037 | 6.83 | 0.41-13.26 | 0.037 |
| +3 vs T0 | 11.27 | 5.66-16.88 | <0.001 | 11.08 | 5.44-16.71 | <0.001 |
| +6 vs T0 | 17.87 | 12.18-23.57 | <0.001 | 17.82 | 12.10-23.54 | <0.001 |
| **Sex** |  |  |  |  |  |  |
| Female | Reference |  |  | Reference |  |  |
| Male vs female | 14.51 | -2.61-31.63 | 0.096 | 17.21 | 0.47-33.95 | 0.044 |
| **Age** |  |  |  |  |  |  |
| Per 10-year increase | -12.82 | -24.59--1.04 | 0.033 | -15.23 | -27.91--2.56 | 0.019 |
| **Education** |  |  |  |  |  |  |
| Complete primary | Reference |  |  | Reference |  |  |
| Less than complete primary vs complete primary | -35.29 | -62.76--7.83 | 0.012 | -31.78 | -59.63--3.94 | 0.025 |
| Complete secondary vs complete primary | 11.64 | -7.72-30.99 | 0.237 | 3.13 | -16.59-22.85 | 0.755 |
| Higher education vs complete primary | -9.03 | -34.97-16.91 | 0.493 | -21.19 | -47.04-4.67 | 0.108 |
| **Marital status** |  |  |  |  |  |  |
| Widowed / separated / divorced | Reference |  |  | - | - | - |
| Single vs widowed/separated/divorced | 1.85 | -26.62-30.32 | 0.898 | - | - | - |
| Married vs widowed/separated/divorced | 3.68 | -26.51-33.87 | 0.810 | - | - | - |
| Cohabiting vs widowed/separated/divorced | 3.44 | -36.19-43.06 | 0.864 | - | - | - |
| **Psychiatric diagnoses** |  |  |  |  |  |  |
| Depressive disorder: present vs absent | 19.17 | 0.68-37.65 | 0.042 | 20.52 | 1.66-39.37 | 0.033 |
| Bipolar disorder: present vs absent | -12.13 | -42.50-18.24 | 0.432 | - | - | - |
| Psychotic disorder: present vs absent | 5.37 | -11.57-22.31 | 0.533 | - | - | - |
| Anxiety/stress-related disorder: present vs absent | 19.33 | -5.97-44.64 | 0.134 | 8.47 | -17.72-34.67 | 0.524 |
| Dementia: present vs absent | 5.39 | -22.10-32.88 | 0.700 | - | - | - |
| **Diagnostic burden** |  |  |  |  |  |  |
| 0-1 categories | Reference |  |  | Reference |  |  |
| 2 vs 0-1 categories | 26.55 | 6.03-47.06 | 0.011 | 32.74 | 12.07-53.41 | 0.002 |
| >=3 vs 0-1 categories | 13.95 | -21.52-49.41 | 0.439 | 19.21 | -16.72-55.15 | 0.293 |
| **Clinical history** |  |  |  |  |  |  |
| Duration of psychiatric illness, per year | -0.19 | -0.75-0.36 | 0.490 | - | - | - |
| Duration of care at INSM, per year | -0.02 | -0.67-0.63 | 0.951 | - | - | - |

Notes: beta = regression coefficient; CI = confidence interval. Individual models included time and one candidate covariate at a time, with a participant-specific random intercept. Entry into the multivariable model was based on the overall covariate p value; for categorical variables with more than two levels, overall association was assessed using a Type III Wald F test with Satterthwaite-approximated degrees of freedom. Covariates with overall $p < 0.20$ in the current analytic dataset were entered into the adjusted model. A hyphen indicates variables that did not meet the screening criterion and were therefore not entered into the adjusted model. T0 is the reference time point. Age is expressed per 10-year increase. Covariate-specific adjusted estimates are exploratory because variable selection was data-driven. The adjusted complete-case model included 212 participants contributing 878 observations.

### Exploratory associations with sociodemographic and clinical characteristics

The individual-model screening step identified sex, age, education, depressive disorder, anxiety or stress-related disorder, and diagnostic burden for inclusion in the multivariable model using the prespecified overall p < 0.20 criterion. In the adjusted exploratory model, male sex was associated with a higher medication score (beta = 17.21; 95% CI 0.47 to 33.95; p = 0.044), whereas each 10-year increase in age was associated with a lower score (beta = -15.23; 95% CI -27.91 to -2.56; p = 0.019). Compared with complete primary education, less than complete primary education was associated with a lower medication score (beta = -31.78; 95% CI -59.63 to -3.94; p = 0.025). Depressive disorder was associated with a higher score (beta = 20.52; 95% CI 1.66 to 39.37; p = 0.033), whereas the adjusted estimate for anxiety or stress-related disorder was imprecise (beta = 8.47; 95% CI -17.72 to 34.67; p = 0.524). Having two psychiatric diagnostic categories, compared with 0-1, was associated with a higher medication score (beta = 32.74; 95% CI 12.07 to 53.41; p = 0.002), while the estimate for three or more categories was imprecise (beta = 19.21; 95% CI -16.72 to 55.15; p = 0.293) (Table 3).

### Exploratory effect modification of the T0-to-+6-month change

The exploratory interaction analysis used a common complete-case sample of 212 participants and 407 observations at T0 or +6 months; 195 participants contributed both time points. Age showed nominal evidence of effect modification: each 10-year increase in age was associated with a 10.94-point greater T0-to-+6-month increase in medication score (95% CI 2.33 to 19.55; global interaction p = 0.013). However, this interaction did not remain statistically significant after Benjamini-Hochberg false-discovery-rate correction (FDR-adjusted p = 0.091). The age model estimated T0-to-+6-month increases of 11.42 points at age 60, 22.36 points at age 70, and 33.30 points at age 80; these estimates are illustrative predictions from the continuous interaction model rather than age thresholds. No other global interaction showed evidence of effect modification after FDR correction: sex (global p = 0.707; FDR p = 0.733), education (p = 0.480; FDR p = 0.671), depressive disorder (p = 0.132; FDR p = 0.461), anxiety or stress-related disorder (p = 0.254; FDR p = 0.561), psychosis (p = 0.321; FDR p = 0.561), and diagnostic burden (p = 0.733; FDR p = 0.733) (Table 4). For psychosis, included a priori, the interaction estimate was -6.34 points (95% CI -18.90 to 6.22), indicating no clear difference in the T0-to-+6-month change between participants with and without psychosis.

**Table 4.** Exploratory effect modification of the T0-to-6-month change in psychotropic medication score.

| Potential effect modifier | Comparison | Interaction beta | 95% CI | Coefficient P | Global interaction P | FDR-adjusted P |
| --- | --- | --- | --- | --- | --- | --- |
| **Sex** | Male vs female | -2.44 | -15.19-10.32 | 0.707 | 0.707 | 0.733 |
| **Age** | Per +10 years | 10.94 | 2.33-19.55 | 0.013 | 0.013 | 0.091 |
| **Education** | Overall interaction | — | — | — | 0.480 | 0.671 |
|  | Less than complete primary vs complete primary | 3.87 | -17.11-24.85 | 0.716 | — | — |
|  | Complete secondary vs complete primary | 2.20 | -12.47-16.87 | 0.768 | — | — |
|  | Higher education vs complete primary | -12.20 | -31.54-7.14 | 0.215 | — | — |
| **Depression** | Present vs absent | 10.70 | -3.24-24.65 | 0.132 | 0.132 | 0.461 |
| **Anxiety** | Present vs absent | 10.71 | -7.77-29.19 | 0.254 | 0.254 | 0.561 |
| **Psychosis** | Present vs absent | -6.34 | -18.90-6.22 | 0.321 | 0.321 | 0.561 |
| **Diagnostic burden** | Overall interaction | — | — | — | 0.733 | 0.733 |
|  | 2 vs 0-1 categories | 6.02 | -9.20-21.24 | 0.437 | — | — |
|  | >=3 vs 0-1 categories | 3.08 | -23.35-29.52 | 0.818 | — | — |

*Notes: beta is the estimated difference in T0-to-+6-month change relative to the stated reference category, or the additional change per 10-year increase in age. Each exploratory model included time, the candidate effect modifier, all main effects for sex, age, education, depressive disorder, anxiety/stress-related disorder, psychosis, and diagnostic burden, plus one time-by-modifier interaction and a participant-specific random intercept. Psychosis was included a priori on clinical/theoretical grounds, independently of the Table 3 screening rule. Global interaction P values are from Type III Wald F tests with Satterthwaite-approximated degrees of freedom. FDR-adjusted P values use the Benjamini-Hochberg procedure across the seven global interaction tests. For education and diagnostic burden, level-specific coefficients are shown beneath the overall interaction row. The common complete-case sample comprised 212 participants and 407 observations at T0 or +6 months; 195 participants contributed both time points. These analyses are exploratory.*

### Sensitivity and robustness analyses

The primary T0-to-+6-month contrast was consistent across the prespecified and post-audit sensitivity analyses. The estimated change was 18.71 points using the alternative medication score definition (95% CI 13.75 to 23.66), 19.49 points after restricting the cohort to participants aged 60 years or older at T0 (95% CI 13.44 to 25.55), 20.28 points after restricting to those aged 60 years or older at the nominal start of follow-up (95% CI 13.58 to 26.99), and 21.89 points using the literal protocol wording of age greater than 60 years at the nominal start of follow-up (95% CI 14.42 to 29.36). Excluding the 10 participants with the most extreme residual patterns yielded an estimate of 17.04 points (95% CI 12.30 to 21.78). All of these estimates remained in the same direction as the primary analysis (Supplementary Table S1).

Participant-level nonparametric cluster bootstrap analysis further supported the primary inference. All 2,000 bootstrap models converged and none was singular; the bootstrap mean contrast was 17.77 points and the percentile 95% CI was 11.77 to 24.06. Visual diagnostics showed departures from ideal Gaussian assumptions, including heavier residual tails and some heteroscedasticity at higher fitted values, but the influence analysis and bootstrap results did not indicate that the primary finding was driven by a small number of participants or by model instability (Supplementary Table S4).

## Discussion

In this retrospective longitudinal cohort, dose-normalized psychotropic medication exposure increased from the last consultation before patients were transferred to the older adult mental health service (T0) to 6 months later, with a mean difference of 17.87 points (95% CI 12.18 to 23.57). The exploratory adjusted estimate was nearly identical at 17.82 points (95% CI 12.10 to 23.54), and the direction and magnitude of the primary contrast were consistent across alternative medication-score definitions, age-restricted analyses, influence analysis, and participant-level bootstrap resampling. This increase, however, should be interpreted within the broader longitudinal trajectory. Estimated mean medication scores declined from 104.9 points at -6 months and 101.6 at -3 months to 94.8 at T0, before increasing to 106.1 at +3 months and 112.7 at +6 months. Thus, the primary T0-to-+6-month contrast might capture an increase from a relative pre-transfer low point rather than evidence of a monotonic escalation beginning around transfer. Exploratory adjusted analyses suggested that average medication-score levels were higher among men, participants with depressive disorder, and those with two diagnostic categories, and lower with older age and less than complete primary education. In separate effect-modification analyses, age was the only characteristic showing evidence of heterogeneity in the T0-to-+6-month change, although this association did not remain statistically significant after correction for multiple comparisons.

Substantial changes in psychotropic prescribing have also been observed around other late-life care transitions. For example, in a population-based study of 250,617 adults aged 65 years or older in Northern Ireland, antipsychotic dispensing increased from 8.2% before entry into residential care to 18.6% after entry, while hypnotic dispensing increased from 14.8% to 26.3% [25]. Similarly, among 322,120 older adults entering residential aged care in Australia, 21.3% received an antipsychotic, 30.5% a benzodiazepine, and 37.9% an antidepressant during the first three months after entry; notably, 45.7% of those receiving an antipsychotic during this period had not received one in the preceding year [26]. In a Norwegian nursing-home cohort of 696 residents, the incidence of major psychotropic drug classes was highest during the first six months after admission, reaching 49.4% for antipsychotics [24].

More directly comparable evidence from specialist outpatient psychiatry also suggests that transitions or structured medication review can be accompanied by substantial treatment reconfiguration rather than a uniform increase or decrease in medication exposure. In a Danish register-based study of 792 older adults referred to secondary outpatient psychiatric care for depression, 25% discontinued antidepressant treatment within 90 days of the first outpatient contact; among the 594 who continued treatment, 47% switched antidepressants. Among patients who remained on the same antidepressant and had recorded dose information, 38% underwent a dose adjustment [32]. A contrasting example comes from Latin America, where deprescribing was an explicit clinical objective. In a quasi-experimental study of 150 adults aged 60 years or older attending a geriatric psychiatry outpatient clinic in Brazil, deprescribing at least one psychotropic medication was recommended for 61.3% of participants and was implemented in 68.5% of those cases [33]. A systematic review of specialist outpatient interventions likewise identified reductions in medication load in four randomized trials, although all included studies were judged at high risk of bias [27].

Taken together, the evidence suggests that psychotropic treatment can change substantially around late-life care transitions and specialist medication review, although the direction and nature of these changes may vary across settings, clinical objectives, drug classes, and prior treatment history. In this context, the increase observed in our cohort may be better understood as a change in overall prescribed psychotropic exposure occurring around transfer between services within the same institution.

Exploratory analyses showed higher average medication-score levels among participants with two recorded diagnostic categories than among those with 0–1 categories, while diagnostic burden was not associated with a differential T0-to-+6-month change. This pattern could reflect the use of complementary psychotropic classes to address more than one psychiatric condition, without necessarily requiring higher doses of individual medications. Regional evidence likewise suggests a substantial burden of psychotropic polypharmacy in psychiatric populations, although these studies have generally measured medication counts rather than dose-normalized exposure. In Argentina, CNS-active polypharmacy among older outpatients was strongly associated with schizophrenia, bipolar disorder, and depressive disorder [21]. At the same institution as the present study, psychotropic and antipsychotic polypharmacy were common among clinically stable outpatients with schizophrenia [22]. Similarly, in a national multicenter sample of 2,475 psychiatric patients in Brazil, psychotropic polypharmacy was present in 85.3%, and multiple psychiatric diagnoses were associated with polypharmacy in both sexes; prevalence was similar in men and women (85.7% vs 84.9%; p > 0.05) [34].

Depressive disorder was also associated with a 20.52-point higher average medication score (95% CI 1.66 to 39.37), consistent with previous evidence linking depression to greater medication burden. In Argentina, depressive disorders were associated with CNS-active polypharmacy (OR 3.50) [21], while a Dutch study of older adults found polypharmacy in 46.9% of participants with depression compared with 19.7% of those without depression; after adjustment, depression remained associated with polypharmacy (OR 1.88; 95% CI 1.02 to 3.48) [35]. Male sex was also associated with a higher average medication score in our cohort (17.21 points; 95% CI 0.47 to 33.95), although previous findings regarding sex have been inconsistent. In the Brazilian PESSOAS study, psychotropic polypharmacy was nearly identical in men and women (85.7% vs 84.9%; p > 0.05) [34], whereas a community-based study of 1,635 older Brazilian adults found greater psychotropic use among women (OR 2.20; 95% CI 1.49 to 3.27) [36].

Regarding education, previous evidence has shown heterogeneous associations between education and medication use. Among older adults with depression in the Netherlands, high educational attainment was associated with lower odds of polypharmacy than low education (adjusted OR 0.43; 95% CI 0.21 to 0.89), whereas among older Costa Rican adults with a self-reported psychiatric diagnosis, psychotropic medication use increased with educational level [37]. In our cohort, differences in average medication scores were observed only between the two lowest educational categories, rather than across the full educational gradient, making this finding difficult to interpret.

Older age was associated with lower average medication-score levels across follow-up. In contrast, each 10-year increase in age was associated with an approximately 11-point greater T0-to-+6-month increase, although this interaction did not remain statistically significant after false-discovery-rate correction. Previous evidence suggests that older individuals may receive lower antipsychotic doses: in 32,062 inpatients with schizophrenia, Zolk et al. found that antipsychotic doses increased until approximately age 40 years and subsequently declined, with a steeper decrease after age 55 years [17]. In a prospective study of stable late-life schizophrenia, Graff-Guerrero et al. found that antipsychotic dose reduction was feasible in most participants and suggested a lower therapeutic dopamine D2/3 receptor occupancy window in older than in younger patients [38]. The greater T0-to-+6-month increase with advancing age is more difficult to explain. Possible explanations include broader treatment reassessment after transfer among older patients, such as treatment of additional symptoms or syndromes with complementary psychotropic agents, or a greater number of medication changes during adaptation to the new service; however, these mechanisms were not directly assessed in our data.

### Strengths and limitations

This study has several strengths. It used repeated routine-care measurements from a specialized older adult mental health service and reconstructed a visit-level medication score from individual prescriptions using explicit dose-parsing and reference-dose rules, with consistent findings across alternative score definitions. The longitudinal mixed-effects model accommodated the unbalanced number of observations across time points while accounting for within-participant correlation. The primary T0-to-+6-month contrast was specified separately from exploratory covariate analyses, and effect modification was evaluated in a distinct set of interaction models using a common analytic sample and false-discovery-rate correction across seven global hypotheses. Robustness was further examined through age-restricted analyses, influence analysis, diagnostic plots, and 2,000 participant-level bootstrap resamples.

Nevertheless, several limitations should frame interpretation. First, this was a retrospective single-cohort study without a contemporaneous comparison group, so the observed trajectory may reflect symptom course, prior undertreatment, treatment optimization, documentation practices, natural within-person fluctuation around the relative low point at T0, or other time-varying clinical factors rather than an effect of service transfer itself. Visit timing was also approximate because observations were drawn from routine care using the eligible consultation closest to each prespecified time point. Missed visits and direct entry into the older adult service resulted in an unbalanced longitudinal dataset, potentially introducing bias if the occurrence or timing of observed visits was related to clinical status, treatment changes, or patterns of service use.

Second, the medication score was an operational measure of dose-normalized prescribed psychotropic exposure rather than a validated measure of prescribing appropriateness, toxicity, or clinical benefit, and no minimal clinically important difference has been established for this score. The observed 17.87-point increase therefore cannot by itself be interpreted as beneficial, harmful, or inappropriate prescribing. Different combinations of drugs and doses may generate similar aggregate scores, and the score should not be interpreted as implying pharmacological equivalence between regimens. Reference-dose definitions also vary across psychotropics, and alternative standardization methods may yield different equivalence estimates [18-18], although the similar primary contrast obtained with an alternative score specification supports the robustness of the direction and approximate magnitude of the association. In addition, the score reflects scheduled psychotropic doses prescribed and documented at eligible INSM visits rather than medication actually obtained or taken. PRN-only prescriptions were excluded, and treatment received outside the INSM or changes occurring between eligible visits may have been incompletely captured.

Third, the analytic cohort reflected actual service transfer rather than a strict age threshold, because transfer to the older adult service was based on clinical and service considerations rather than a fixed age cutoff. Age-restricted sensitivity analyses yielded estimates in the same direction, reducing concern that the primary finding was driven by participants younger than 60 years. Finally, visual diagnostics showed heavier residual tails and some heteroscedasticity, and the exploratory multivariable model used data-driven variable selection. Stable influence analyses and participant-level bootstrap estimates support the robustness of the primary contrast, but do not eliminate residual model misspecification, selection, measurement, or confounding.

### Clinical and health-system implications

Clinically, transfer to a specialized older adult service may provide an opportunity for structured medication review focused on current indication and target symptoms, treatment response and tolerability, drug-drug interactions and geriatric vulnerabilities, and whether continuation, dose adjustment, substitution, or deprescribing remains justified. Medication amount, however, should not be equated with prescribing appropriateness. Potentially inappropriate prescribing has been reported among older adults in Peru using STOPP/START criteria [23], while in Colombia 37.1% of 11,372 people with dementia received an antipsychotic, a prescribing pattern considered potentially inappropriate in that clinical context [39]. Together, these studies support individualized medication review during service transitions in older adults.

From a health-system perspective, transfer between services may also provide a useful checkpoint for medication continuity and reconciliation, particularly because many participants had received care at the INSM for years before transfer and were changing treating services rather than initiating psychiatric care. Prescribing decisions may therefore accumulate across clinicians and services, increasing the importance of preserving information on treatment indication, response, adverse effects, and previous medication changes across handoffs. In a systematic review and meta-analysis of 24 randomized studies including 17,664 older adults, interventions incorporating medication reconciliation were associated with a lower risk of hospital readmission (RR 0.88; 95% CI 0.81 to 0.96) [40]. Although those studies predominantly involved hospital discharge, they support evaluating medication reconciliation, structured information transfer, and coordinated referral and counter-referral processes during transitions between psychiatric services.

### Conclusions and future research

Future research should combine medication trajectories with repeated measures of symptoms, cognition, functioning, adverse effects, hospitalization, quality of life, and adherence to determine whether changes in prescribed psychotropic exposure correspond to clinically meaningful outcomes. Comparative designs including patients who remain in general adult services or follow alternative care pathways would help distinguish changes associated with service transfer from underlying temporal trends. Future studies should also characterize medication changes at the drug and class level—including initiation, discontinuation, switching, and dose adjustment—and incorporate validated measures of prescribing appropriateness, anticholinergic burden, and sedative burden. Prospective designs with prespecified assessment windows and better characterization of continuity of care, medication availability, caregiver involvement, and referral pathways would further strengthen interpretation.

In this cohort, dose-normalized prescribed psychotropic exposure increased from a relative pre-transfer low point at T0 to 6 months after transfer, with similar estimates across multiple sensitivity analyses. The findings indicate a consistent temporal increase in documented prescribed exposure around service transfer, while the clinical meaning and mechanisms underlying this change remain to be established.

## Declarations

## Acknowledgments

The authors thank the Instituto Nacional de Salud Mental Honorio Delgado–Hideyo Noguchi for institutional support, and the clinical, nursing, and administrative staff of the older adult service of the DEIDAE de Adultos y Adultos Mayores for their work in the care and documentation of the patients on whom this study is based. The authors also thank the staff of the medical records and information systems units for their assistance during data extraction.

## Use of generative artificial intelligence

Generative artificial intelligence tools (OpenAI ChatGPT) were used during the development of this study to assist with drafting and debugging analytic code, organization and review of literature, consistency checks across statistical outputs, tables, and manuscript text, methodological and reporting review, and language and editorial refinement. Part of this work was conducted using a structured AI-assisted review workflow developed by one of the authors (Paulo Ruiz-Grosso), designed to organize methodological, statistical, reporting, and editorial checks during manuscript development. This workflow was used as an internal aid and was not treated as an independently validated methodological instrument. All statistical analyses were executed on the study data and reviewed by the authors. The authors independently verified the cited literature, numerical results, interpretations, and final manuscript text, made all substantive scientific and methodological decisions, and take full responsibility for the content of the manuscript.

## Data availability statement

The data are not publicly available because they derive from sensitive clinical records. De-identified analytic data may be available from the corresponding author upon reasonable request and subject to institutional and ethics approvals.

