## Supplemental Tables for "Longitudinal changes in dose-normalized psychotropic medication exposure around transfer to a specialized older adult mental health service: a retrospective longitudinal study in Peru"

### Supplementary Table S1. Sensitivity and robustness analyses for the T0-to-6-month contrast

| Analysis | N | Estimate | 95% CI | P value | Role |
|---|---|---|---|---|---|
| Primary analysis | 213 | 17.87 | 12.18 to 23.57 | <0.001 | Primary |
| Alternative medication-score definition | 213 | 18.71 | 13.75 to 23.66 | <0.001 | Sensitivity |
| Age restricted: ≥60 years at T0 | 192 | 19.49 | 13.44 to 25.55 | <0.001 | Sensitivity |
| Age restricted: ≥60 years at nominal follow-up start (T0−6 months) | 165 | 20.28 | 13.58 to 26.99 | <0.001 | Sensitivity |
| Age restricted: >60 years at nominal follow-up start (literal protocol wording) | 133 | 21.89 | 14.42 to 29.36 | <0.001 | Sensitivity |
| Influence sensitivity: 10 participants with extreme residuals excluded | 203 | 17.04 | 12.30 to 21.78 | <0.001 | Sensitivity |
| Adjusted complete-case model | 212 | 17.82 | 12.10 to 23.54 | <0.001 | Exploratory adjusted robustness |
| Participant-level cluster bootstrap, 2,000 resamples | 213 | 17.87 | 11.77 to 24.06 | — | Bootstrap robustness |

Notes: The primary row is included as an anchor. Age-restricted analyses evaluate alternative operationalizations of the protocol age wording. The bootstrap row reports the participant-level percentile 95% CI from 2,000 resamples; all 2,000 models converged and none was singular.

### Supplementary Table S2. Medication- and formulation-specific reference upper doses used to construct the dose-normalized psychotropic medication score

| Medication / formulation | Unit / interval | Primary reference upper dose | Alternative reference used in sensitivity analysis | Primary source used in analytic audit | Primary source URL | Supporting source | Supporting source URL |
|---|---|---|---|---|---|---|---|
| Sertraline, oral | mg/day | 200 | — | Drugs.com | https://www.drugs.com/dosage/sertraline.html | | |
| Escitalopram, oral | mg/day | 20 | — | Drugs.com | https://www.drugs.com/dosage/escitalopram.html | | |
| Paroxetine, oral | mg/day | 40 | 60 | FDA/label | | Drugs.com | https://www.drugs.com/dosage/paroxetine.html |
| Mirtazapine, oral | mg/day | 45 | — | Drugs.com | https://www.drugs.com/dosage/mirtazapine.html | | |
| Fluoxetine, oral | mg/day | 80 | — | Drugs.com | https://www.drugs.com/dosage/fluoxetine.html | | |
| Clomipramine, oral | mg/day | 250 | — | Drugs.com | https://www.drugs.com/dosage/clomipramine.html | | |
| Amitriptyline, oral | mg/day | 150 | 300 | Drugs.com | https://www.drugs.com/dosage/amitriptyline.html | | |
| Haloperidol, oral | mg/day | 20 | 100 | eMC SmPC haloperidol oral | https://www.medicines.org.uk/emc/product/10907/smpc | | |
| Risperidone, oral | mg/day | 16 | — | Drugs.com | https://www.drugs.com/dosage/risperidone.html | | |
| Sulpiride, oral | mg/day | 1600 | 2400 | AEMPS-CIMA Dogmatil Fuerte 200 mg | https://cima.aemps.es/cima/dochtml/ft/48558/FT_48558.html | WHO ATC/DDD | |
| Clozapine, oral | mg/day | 900 | — | Drugs.com | https://www.drugs.com/dosage/clozapine.html | | |
| Quetiapine, oral | mg/day | 800 | 750 | Drugs.com | https://www.drugs.com/dosage/quetiapine.html | | |

### Supplementary Table S2. Medication- and formulation-specific reference upper doses used to construct the dose-normalized psychotropic medication score (continued)

| Medication / formulation | Unit / interval | Primary reference upper dose | Alternative reference used in sensitivity analysis | Primary source used in analytic audit | Primary source URL | Supporting source | Supporting source URL |
|---|---|---|---|---|---|---|---|
| Olanzapine, oral | mg/day | 20 | — | Drugs.com | https://www.drugs.com/dosage/olanzapine.html | | |
| Aripiprazole, oral | mg/day | 30 | — | Drugs.com | https://www.drugs.com/dosage/aripiprazole.html | | |
| Trifluoperazine, oral | mg/day | 40 | — | Drugs.com | https://www.drugs.com/dosage/trifluoperazine.html | | |
| Levomepromazine, oral | mg/day | 400 | 1000 | Base de Donnees Publique des Medicaments (Francia) | https://base-donnees-publique.medicaments.gouv.fr/medicament/67348996/extrait | eMC SmPC Levomepromazine | https://www.medicines.org.uk/emc/product/11912/smpc |
| Chlorpromazine, oral | mg/day | 300 | 1000 | eMC SmPC chlorpromazine | | | |
| Haloperidol decanoate | mg/month | 300 | 450 | EMA Haldol Decanoate Annex III | https://www.ema.europa.eu/en/documents/referral/haldol-decanoate-article-30-annex-iii_en.pdf | EMA Assessment Report | https://www.ema.europa.eu/en/documents/referral/haldol-decanoate-article-30-assessment-report_en.pdf |
| Fluphenazine decanoate | mg/month | 100 | — | DailyMed Fluphenazine Decanoate | https://dailymed.nlm.nih.gov/dailymed/drugInfo.cfm?setid=1967dc24-f2a5-4095-baa9-a9d1c5410311 | | |
| Valproate | mg/day | 2000 | 3000 | Young et al. GERI-BD 2017 | https://pmc.ncbi.nlm.nih.gov/articles/PMC6214451/ | Drugs.com | https://www.drugs.com/dosage/valproic-acid.html |
| Lamotrigine | mg/day | 200 | 400 | Lamictal prescribing information / Drugs.com | https://www.drugs.com/pro/lamictal.html | | |
| Phenobarbital | mg/day | 400 | — | Drugs.com | https://www.drugs.com/dosage/phenobarbital.html | | |
| Carbamazepine | mg/day | 1600 | — | Drugs.com | https://www.drugs.com/dosage/carbamazepine.html | | |
| Gabapentin | mg/day | 3600 | — | Drugs.com | https://www.drugs.com/dosage/gabapentin.html | | |

### Supplementary Table S2. Medication- and formulation-specific reference upper doses used to construct the dose-normalized psychotropic medication score (continued)

| Medication / formulation | Unit / interval | Primary reference upper dose | Alternative reference used in sensitivity analysis | Primary source used in analytic audit | Primary source URL | Supporting source | Supporting source URL |
|---|---|---|---|---|---|---|---|
| Phenytoin | mg/day | 600 | — | Drugs.com | https://www.drugs.com/dosage/phenytoin.html | | |
| Lithium | mg/day | 1200 | 1800 | Drugs.com | https://www.drugs.com/dosage/lithium.html | | |
| Clonazepam | mg/day | 4 | 20 | Drugs.com | https://www.drugs.com/monograph/clonazepam.html | | |
| Diazepam | mg/day | 30 | 40 | eMC SmPC diazepam | https://www.medicines.org.uk/emc/product/101911/smpc | Drugs.com | https://www.drugs.com/dosage/diazepam.html |
| Alprazolam | mg/day | 4 | 10 | Drugs.com | https://www.drugs.com/dosage/alprazolam.html | | |
| Bromazepam | mg/day | 18 | 60 | Fuentes regulatorias internacionales | | | |
| Zolpidem, immediate release | mg/day | 10 | 5 | DIGEMID Peru, ficha tecnica zolpidem 10 mg (2021) | https://www.digemid.minsa.gob.pe/Archivos/FichasTecnicas/EspecialidadesFarmaceuticas/2021/EE10291_FT_V01.pdf | | |
| Biperiden | mg/day | 16 | — | Drugs.com | https://www.drugs.com/dosage/biperiden.html | | |
| Memantine, immediate release | mg/day | 20 | 28 | Drugs.com | https://www.drugs.com/dosage/memantine.html | | |
| Donepezil | mg/day | 23 | — | Drugs.com | https://www.drugs.com/dosage/donepezil.html | | |
| Modafinil | mg/day | 200 | 400 | DailyMed / Drugs.com | https://www.drugs.com/dosage/modafinil.html | | |

Notes: Medication contribution = (scheduled prescribed dose / reference upper dose) × 100; visit score = sum of medication-specific contributions. PRN-only and STAT prescriptions did not contribute to the scheduled score. A dash indicates that the primary reference was retained in the global sensitivity specification. These are operational analytic references, not claims of pharmacological equivalence or prescribing appropriateness.

### Supplementary Table S3. Tukey-adjusted pairwise comparisons of estimated marginal medication-score means across time points

| Contrast (first − second) | Mean difference | SE | df | 95% CI | Tukey-adjusted P value |
|---|---|---|---|---|---|
| −6 months vs −3 months | 3.26 | 3.64 | 668.9 | -6.68 to 13.21 | 0.8979 |
| −6 months vs T0 | 10.06 | 3.42 | 673.2 | 0.71 to 19.42 | 0.0278 |
| −6 months vs +3 months | -1.21 | 3.43 | 672.8 | -10.58 to 8.17 | 0.9967 |
| −6 months vs +6 months | -7.81 | 3.45 | 672.6 | -17.26 to 1.64 | 0.1593 |
| −3 months vs T0 | 6.8 | 3.25 | 672.3 | -2.09 to 15.69 | 0.2247 |
| −3 months vs +3 months | -4.47 | 3.26 | 671.8 | -13.38 to 4.44 | 0.6455 |
| −3 months vs +6 months | -11.07 | 3.29 | 672.1 | -20.08 to -2.06 | 0.0073 |
| T0 vs +3 months | -11.27 | 2.86 | 666.9 | -19.09 to -3.46 | <0.001 |
| T0 vs +6 months | -17.87 | 2.9 | 667.7 | -25.80 to -9.94 | <0.001 |
| +3 months vs +6 months | -6.6 | 2.92 | 668.4 | -14.59 to 1.39 | 0.1594 |

Notes: Contrasts are from the validated primary linear mixed-effects model and estimated marginal means. The sign of the estimate follows first time point minus second time point. P values are Tukey-adjusted across the 10 pairwise comparisons. These supplementary comparisons are distinct from the prespecified +6-month versus T0 primary contrast, for which the manuscript reports the unadjusted two-sided P value.

### Supplementary Table S4. Model diagnostics and inferential robustness

| Diagnostic / analysis | Finding | Quantitative result | Status | Interpretation |
|---|---|---|---|---|
| Residuals vs fitted | Heteroscedasticity more apparent at higher fitted values | Visual diagnostic | Observed | Gaussian homoscedasticity is imperfect; no major curvature was identified. |
| Residual Q-Q | Heavy-tail departures | Visual diagnostic | Observed | Motivated robustness assessment rather than transformation of the primary outcome. |
| Random-intercept Q-Q | Departure from normality / asymmetry | Visual diagnostic | Observed | Random-effects normality is imperfect. |
| Influence sensitivity | Primary contrast stable after excluding 10 extreme-residual participants | Estimate 17.04; 95% CI 12.30 to 21.78; N=203 | Robust | No single influential-participant pattern explained the primary finding. |
| Participant-level cluster bootstrap | 2,000 of 2,000 resamples converged; 0 singular fits | Original 17.87; bootstrap mean 17.77; percentile 95% CI 11.77 to 24.06 | PASS | Participant-level resampling supports the direction and magnitude of the primary inference. |

Notes: Overall diagnostic assessment: model assumptions are not perfect, particularly in distributional tails and at higher fitted values, but influence analyses and participant-level cluster bootstrap results support retaining the prespecified primary linear mixed-effects model without outcome transformation or case deletion.
